# Evaluating large language model workflows in clinical decision support: referral, triage, and diagnosis

**DOI:** 10.1101/2024.09.27.24314505

**Authors:** Farieda Gaber, Maqsood Shaik, Vedran Franke, Altuna Akalin

**Affiliations:** Berlin Institute for Medical Systems Biology (BIMSB), Max Delbrück Center for Molecular Medicine, Berlin, Germany

**Author notes:** These authors contributed equally to this work (Farieda Gaber, Maqsood Shaik).

## Abstract

Accurate medical decision-making is critical for both patients and clinicians. Patients often struggle to interpret their symptoms, determine their severity, and select the right specialist. Simultaneously, clinicians face challenges in integrating complex patient data to make timely, accurate diagnoses. Recent advances in large language models (LLMs) offer the potential to bridge this gap by supporting decision-making for both patients and healthcare providers. In this study, we benchmark multiple LLM versions and an LLM-based workflow incorporating retrieval-augmented generation (RAG) on a curated dataset of 2,000 medical cases derived from the Medical Information Mart for Intensive Care database. Our findings show that these LLMs are capable of providing personalized insights into likely diagnoses, suggesting appropriate specialists, and assessing urgent care needs. These models may also support clinicians in refining diagnoses and decision-making, offering a promising approach to improving patient outcomes and streamlining healthcare delivery.

## Introduction

Clinical decision-making is a fundamentally complex process that relies on clinicians applying their knowledge and experience^1^ while considering numerous factors and integrating vast amounts of data to assess patient symptoms, determine the severity of their condition, and choose the most appropriate next steps. This process typically involves combining information from various sources, such as symptoms, vital signs, patient medical history, and various examinations, to arrive at an accurate and timely diagnosis. The ability to correctly interpret this information and make well-founded decisions is crucial for improving patient outcomes. In a saturated healthcare system with increasing amounts and complexity of patient data, fewer healthcare professionals face the challenge of meeting increasing patient demands for fast, accurate, and personalized care. Especially in high-pressure environments like emergency departments, the fast pace and complexity of decision-making can contribute to delays or errors in triaging, diagnosis and treatment, ultimately leading to suboptimal care.

Recent advancements in large language models (LLMs) have demonstrated significant potential to transform various fields, including clinical decision-support ^2,3^. While LLMs have shown promise in structured environments, such as medical licensing exams and clinical vignettes^4,5^, their application in real-world, open-ended clinical scenarios remains an emerging area of research. Powerful models could increase diagnostic accuracy, optimize triage processes and improve patient management. For example, LLMs could assist in prioritizing patients based on symptoms and vital signs, distinguishing between urgent and non-urgent cases, thereby reducing waiting times and improving care delivery. This capability is especially crucial in emergency departments (EDs), where accurate triage (level of severity of a patient’s condition) assessment is vital for patient prioritization. Errors in this process—whether under-triage (assigning lower urgency than needed) or over-triage (assigning higher urgency)— significantly impact patient outcomes and resource allocation. Trauma systems have set the goal to minimize under triage and accept a higher rate of over triage to reduce mortality rate caused by under triage, with goals set at ≤5% and ≤35%^6^, respectively. A review of field triage performance showed 14% to 34% under triaged cases across all ages^7^, which can result in delayed treatment for patients requiring immediate care, potentially worsening their outcomes. On the other hand, over-triage rates were shown to be between 12% and 31%^7^, leading to the waste of critical resources and increased waiting times for other patients. In this context, LLMs might mitigate both under-triage and over-triage, thereby improving resource allocation and overall patient outcomes.

Beyond assisting clinicians, LLMs could help patients manage their own healthcare decisions. These models have the potential to guide patients in interpreting their symptoms, recommend appropriate specialists and determine the best course of action. However, while the capabilities of LLMs are promising, their real-world application in dynamic and unstructured clinical environments remains an area of active research and development. While the scope of current LLM research in healthcare focuses on diagnosing specific diseases or targeting particular medical specialties, which are necessary and hold significant promise^8–14^, it misses the broader task of predicting diagnoses to support comprehensive clinical decision-making in more general, fast-paced environments. Other studies employ models that are required to choose a diagnosis from a simplified set of binary or multiple-choice options testing human competencies within particular domains^15–18^ which reduces the complexity of real-world clinical decision-making. In practice, clinicians are frequently faced with vague or unclear symptoms, incomplete information, and unlike in controlled studies, they do not have the convenience of selecting from multiple-choice options. Instead, they must rely on their clinical judgment and experience to navigate uncertainty and arrive at a diagnosis.

In this study, we aimed to benchmark multiple LLM workflows on their ability to predict key aspects of clinical care: triage level in the form of the Emergency Severity Index (ESI)^19^, patient to medical specialty referral, and diagnosis based on symptoms (also referred to as history of present illness), patient information and initial vitals. The workflow is illustrated in Figure 1. Using a dataset of 2,000 real-world cases from the Medical Information Mart for Intensive Care (MIMIC-IV) database^20–22^, we evaluate the performance of several LLMs, specifically multiple versions of the Claude family^23,24^, as well as a RAG agentic workflow designed to mimic the clinical decision-making process.

**Figure 1:**
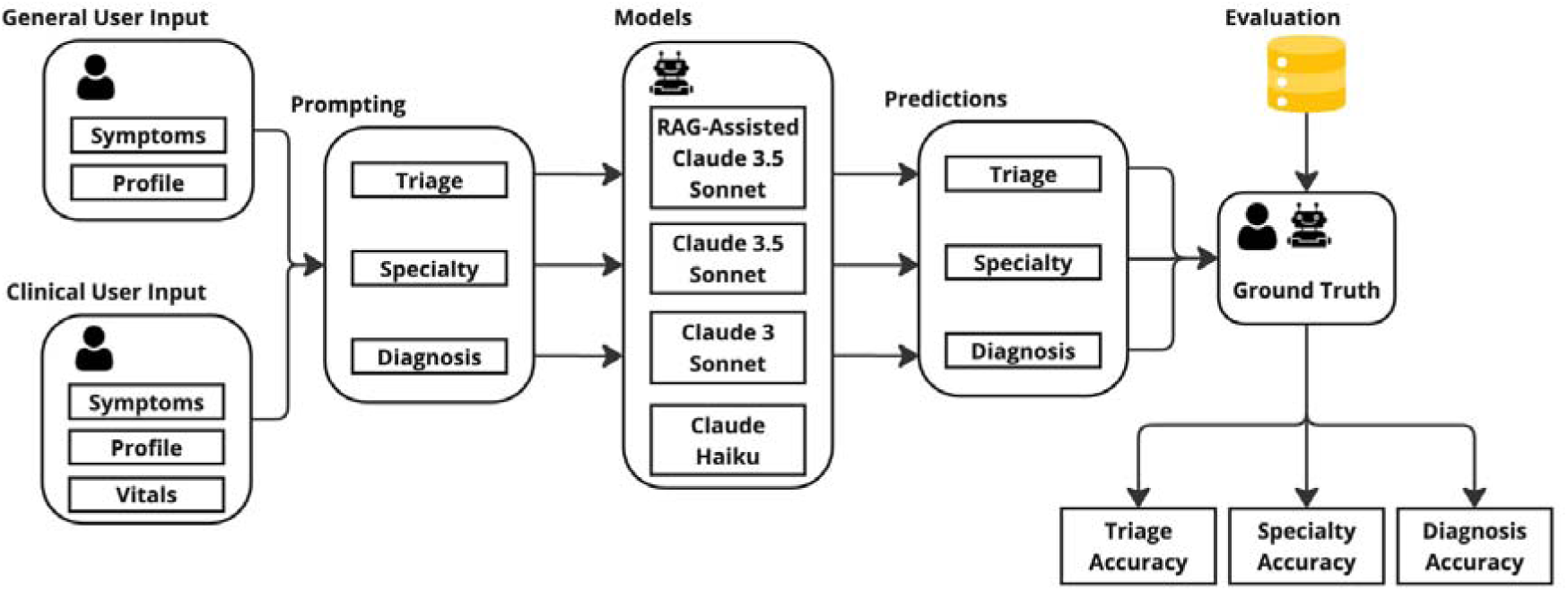
LLM Evaluation Workflow: referral, triage, diagnosis.

This paper systematically evaluates the potential and limitations of these models in supporting clinicians with complex decision-making, showing promising results in their ability to assist effectively. With the increasing digitization of healthcare, the integration of AI-powered tools presents a promising opportunity to enhance clinical workflows and streamline patient-centered care. Such advancements will benefit both clinicians and patients.

## Results

### Curated MIMIC-ED Dataset and Model Evaluation Approach

We created a curated dataset using the fully de-identified MIMIC-IV ED dataset^22,25^, consisting of electronic health records, together with the MIMIC-IV Notes^22,26^ to simulate clinical decision-making in an emergency department setting. Both datasets are modules from MIMIC-IV^20,22^. Details about the dataset and the preprocessing can be found in the Methods: Data Preprocessing. From the processed data, we extracted 2,000 medical cases covering a wide range of medical conditions. Figure 2a) displays the distribution of triage levels in the emergency department (ED), while Figure 2b) shows the specialties managing these cases, occurring more than 30 times. As expected in the ED, there were few triage level 4 and no triage level 5 cases (less severe), with most classified as triage level 3, followed by triage level 2, and a smaller number as triage level 1. This dataset has the advantage of not being directly publicly available, which makes it ideal for evaluating LLMs that otherwise tend to use publicly available test sets as part of their training data.

**Figure 2:**
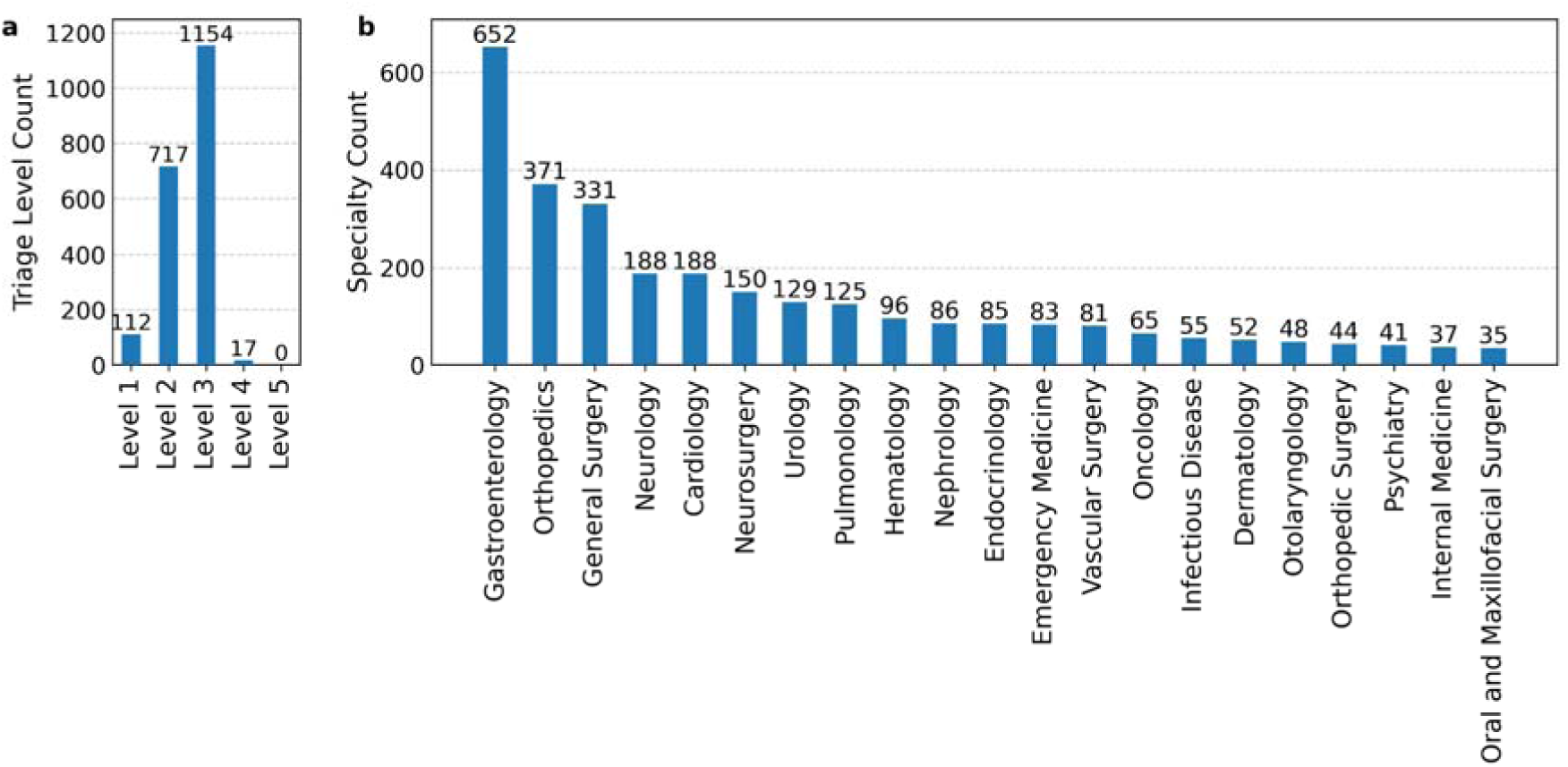
a) shows the quantities of the triage levels and b) shows the quantities of the most frequent specialties.

**Figure 3:**
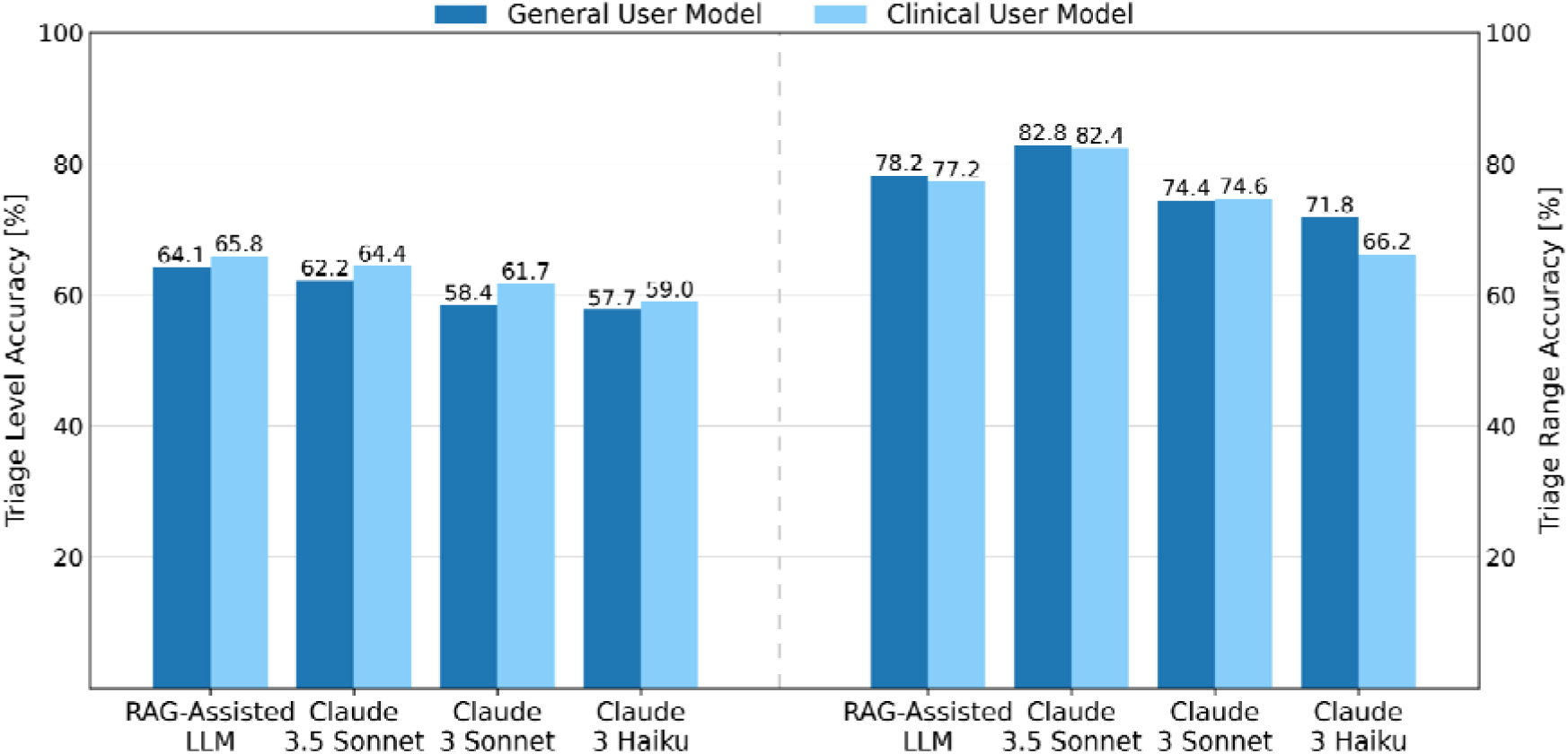
Performance as accuracy [%] on triage level with exact match evaluation on the left and range evaluation on the right for both model types.

### Model Selection and RAG-Assisted LLM

We tested three models from the Claude family - Claude 3.5 Sonnet, Claude 3 Sonnet, and Claude 3 Haiku - due to their superior performance across multiple benchmarks, excelling in contextual understanding, efficiency, and handling specialized queries^23,24^ (see Methods: Model Selection for details).

Additionally, we developed a RAG-assisted LLM The model is built on Claude 3.5 Sonnet and incorporates a multi-step process where the LLM plays a key role in refining and enhancing query processing and answer generation. The workflow starts with a query decomposition, breaking down the patient’s query into smaller queries, mimicking what clinicians or curious patients might do. These individual queries are then searched in a semantic database to find matching PubMed abstracts, which provides an additional layer of accountability and allows users to easily verify the model’s results. The retrieved abstracts are used alongside the original query to generate a response from the LLM, with the expectation that these abstracts prevent the LLM from “hallucinating” information. This process is known as Retrieval-Augmented Generation. To simulate a more advanced workflow, the system incorporates additional loops of critique, refinement, and retrieval, all using LLMs.

Due to privacy regulations surrounding the MIMIC-IV dataset, which prohibit its use with external application programming interfaces (APIs) like those provided by OpenAI (e.g., GPT-4), we utilized AWS Privatelink to privately connect to the Claude models supported by AWS services. More details are provided in the Methods: Model Selection.

For each model we differentiated between two user types: general users, typically patients who provide only personal information and symptoms (referred to as the ‘history of present illness’ in the dataset), and clinicians in the ED, who can additionally retrieve initial clinical data, such as temperature, heart rate, respiratory rate, oxygen saturation, and blood pressure. While we recognize that making a definitive diagnosis requires further input, such as physical exams or laboratory tests, our approach seeks to replicate the decision-making process both for patients feeling ill at home and those arriving at the ED. This distinction allowed us to explore the capabilities of LLMs in both home settings, where users report symptoms, and ED settings, where preliminary clinical data is available.

### LLM performance in triage

In the context of emergency care, triage or acuity as it is mentioned in the MIMIC-IV-ED dataset refers to the severity of a patient’s condition and is commonly assessed using the Emergency Severity Index (ESI)^19^. This standardized triage tool classifies patients into five levels based on the urgency of their treatment needs, allowing healthcare providers to prioritize care more effectively. The levels range from ESI 1, which indicates patients requiring immediate life-saving interventions, to ESI 5, which represents cases where treatment can be safely delayed. The description for each level can be found in the Supplementary Information in Table S1. This classification system plays a crucial role in emergency department operations, helping clinicians to allocate resources efficiently and address critical cases with minimal delay.

In our study, we assess the model’s capabilities to predict patient triage level for two user scenarios: a general user providing only symptom-based information and a clinician with additional access to initial clinical data. This evaluation aims to determine whether the models can be effectively integrated into the decision-making process to assist in triaging patients in real-time. The specific prompting details used for these cases can be found in the Methods: Prompts.

The results were assessed based once on exact match accuracy, where the predicted triage level matched the actual value, and a triage range accuracy, where predictions were considered correct if they were exactly or only one triage level higher than the actual level, except for the triage level 1, which has to be predicted as 1. Models incorporating vital signs generally performed better in predicting the triage level than those using symptoms alone. RAG-Assisted LLM showed the highest exact match accuracy in both conditions. The addition of clinical data had a modest but positive effect on performance across all models and more recent models outperformed simpler ones.

Under the triage range accuracy metric, Claude 3.5 Sonnet outperformed all other models. All the results are presented in Table 1. Figure 4 presents the confusion matrices for the two models that performed best in each accuracy evaluation. These matrices provide additional insight into the models’ behavior. It is important to note that while no model achieved high accuracy in predicting the most severe triage levels, none of the models confused the most critical cases with the least serious ones, and vice versa. This is a crucial finding, as it indicates that the models had difficulty accurately predicting cases at the extreme ends of triage severity, but they consistently recognized the difference between life-threatening cases and those of lower urgency.

**Figure 4:**
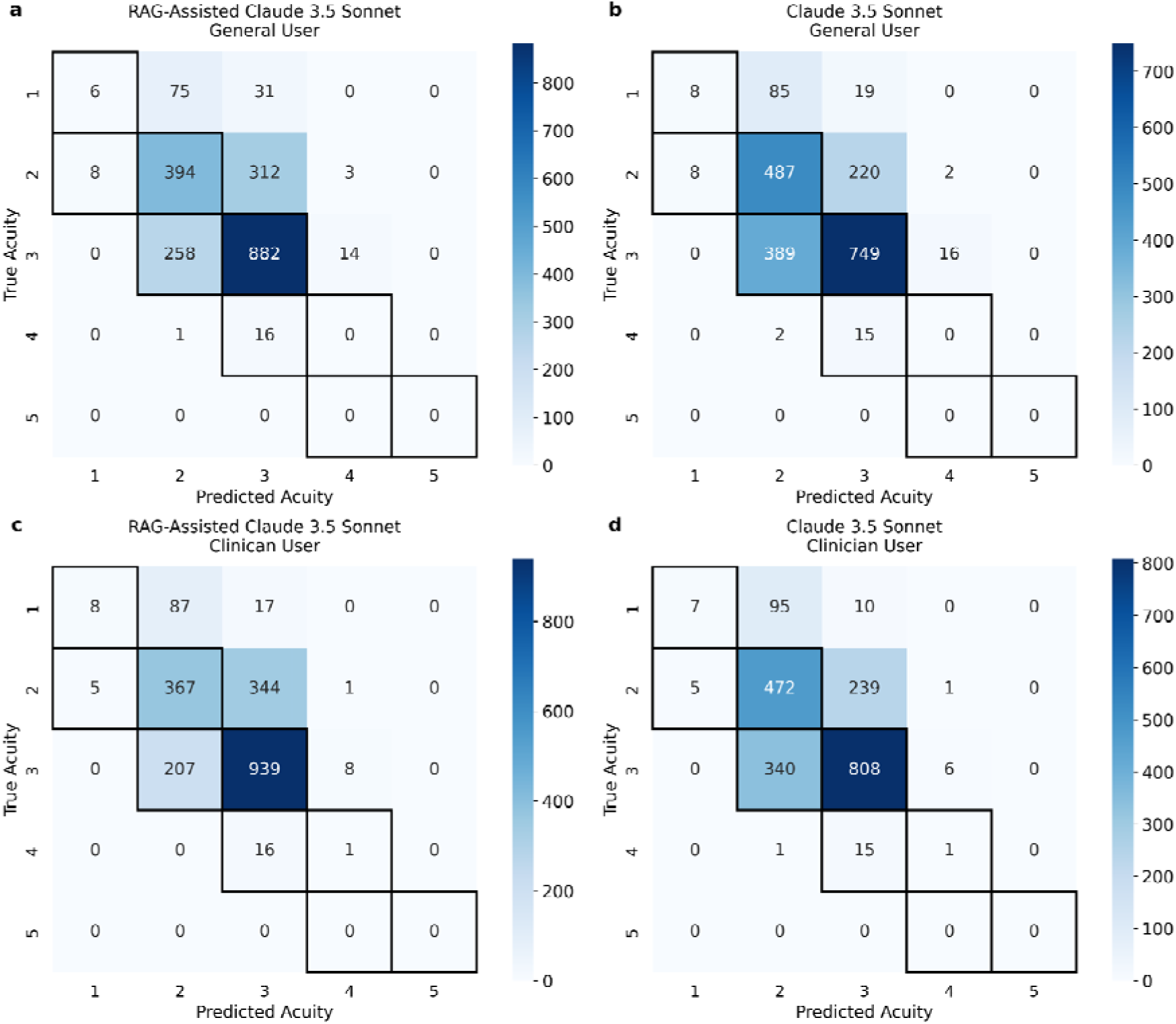
Confusion matrices for the two best models a) and b) for the general user setting and c) and d) for the clinical user setting with the diagonal values counted as correct predictions for the exact match evaluation and the marked predictions counted as correct values for the triage range evaluation

**Table 1:**
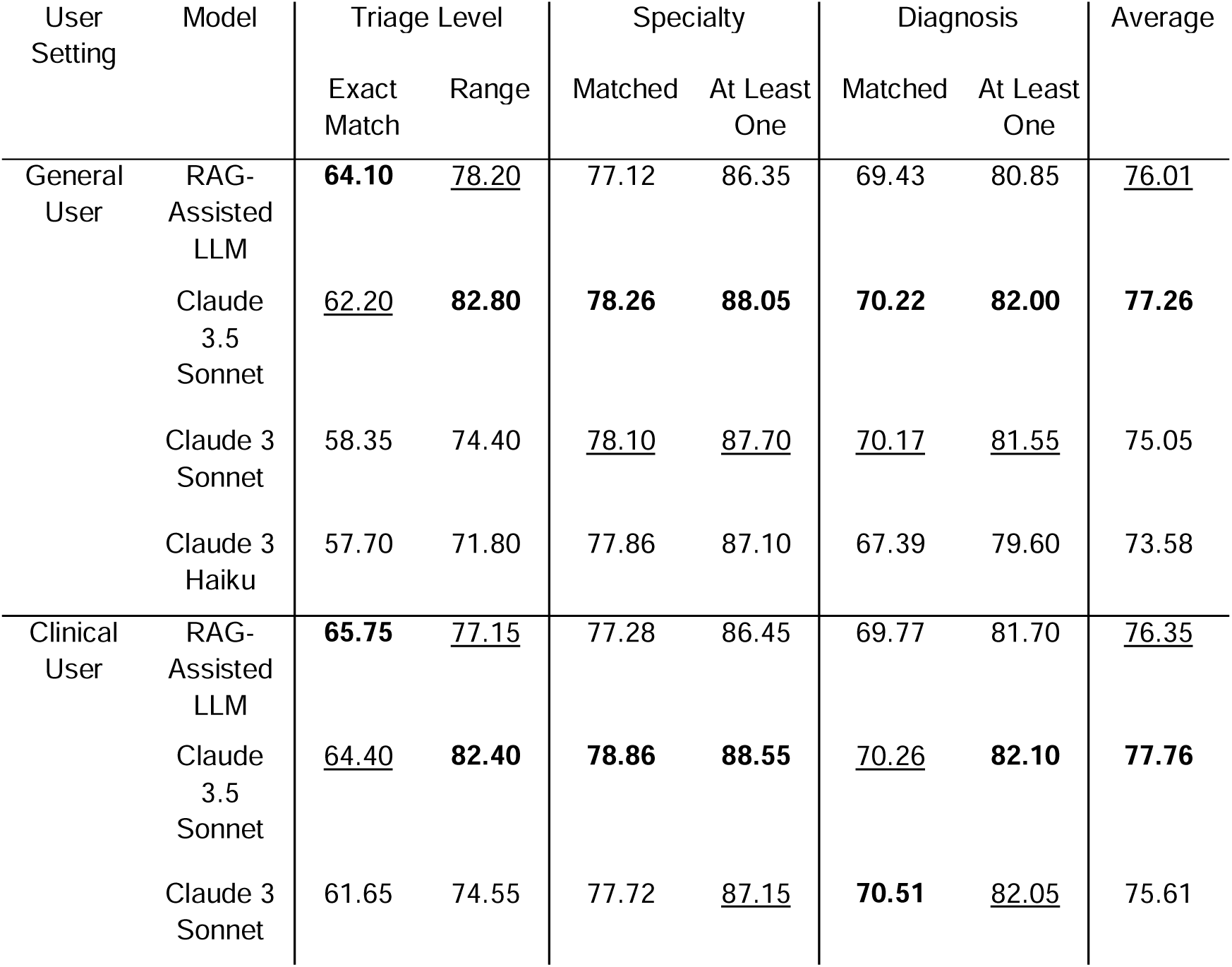
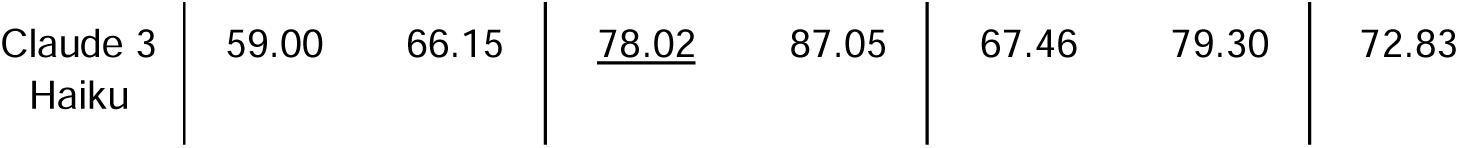
Performance as accuracy [%] on all tasks and with all evaluation methods. Bold marks the best model in each model group and underlined the second best.

| User Setting | Model | Triage Level |  | Specialty |  | Diagnosis |  | Average |
| --- | --- | --- | --- | --- | --- | --- | --- | --- |
|  |  | Exact Match | Range | Matched | At Least One | Matched | At Least One |  |
| General User | RAG-Assisted LLM | **64.10** | <u>78.20</u> | 77.12 | 86.35 | 69.43 | 80.85 | <u>76.01</u> |
|  | Claude 3.5 Sonnet | <u>62.20</u> | **82.80** | **78.26** | **88.05** | **70.22** | **82.00** | **77.26** |
|  | Claude 3 Sonnet | 58.35 | 74.40 | <u>78.10</u> | <u>87.70</u> | <u>70.17</u> | <u>81.55</u> | 75.05 |
|  | Claude 3 Haiku | 57.70 | 71.80 | 77.86 | 87.10 | 67.39 | 79.60 | 73.58 |
| Clinical User | RAG-Assisted LLM | **65.75** | <u>77.15</u> | 77.28 | 86.45 | 69.77 | 81.70 | <u>76.35</u> |
|  | Claude 3.5 Sonnet | <u>64.40</u> | **82.40** | **78.86** | **88.55** | <u>70.26</u> | **82.10** | **77.76** |
|  | Claude 3 Sonnet | 61.65 | 74.55 | 77.72 | <u>87.15</u> | **70.51** | <u>82.05</u> | 75.61 |

|  |  |  |  |  |  |  |  |
| --- | --- | --- | --- | --- | --- | --- | --- |
| Claude 3<br>Haiku | 59.00 | 66.15 | <u>78.02</u> | 87.05 | 67.46 | 79.30 | 72.83 |

The improvement between the general user and clinical user models can be observed in Table 2, which shows a performance increase in triage level prediction across all models. This highlights how the LLM’s predictions improve when provided with more detailed information, similar to how a clinician makes more accurate decisions when given initial vitals. However, this improvement is not as apparent in the triage range evaluation. When the model misclassified the triage level, it is usually within the range of one level more severe. A slight decline in triage range accuracy was noted across most cases, except for Claude 3 Haiku, which struggled strongly to process the additional information from the initial vitals effectively.

**Table 2:** Performance improvement for each model from general user to clinical user setting.

| Model | Triage Level |  | Specialty |  | Diagnosis |  | Average |
| --- | --- | --- | --- | --- | --- | --- | --- |
|  | Exact Match | Range | Matched | At Least One | Matched | At Least One |  |
| RAG-Assisted LLM | 1.65 | -1.05 | 0.16 | 0.10 | 0.34 | 0.85 | 0.34 |
| Claude 3.5 Sonnet | 2.20 | -0.40 | 0.60 | 0.50 | 0.04 | 0.10 | 0.51 |
| Claude 3 Sonnet | 3.30 | 0.15 | -0.38 | -0.55 | 0.34 | 0.50 | 0.56 |
| Claude 3 Haiku | 1.30 | -5.65 | 0.16 | -0.05 | 0.07 | -0.30 | -0.75 |

### Predicting appropriate medical specialty referrals from patient data

We aimed to evaluate whether LLMs can assist in the specialty referral process. Accurate identification of the appropriate specialty for a patient is critical in ensuring they receive the most effective and timely treatment, which also reduces healthcare costs by minimizing unnecessary referrals. Since the MIMIC-IV-ED and MIMIC-IV-Notes datasets don’t contain information on the medical specialist each patient visited, we used Claude 3.5 Sonnet to create a ground truth by predicting the most likely specialist for each of the diagnoses of each patient. More details on this process and the used prompt can be found in Methods: Prompts.

We evaluated the ability of LLMs to predict specialties in our two scenarios, the general user and clinical user models. For each scenario, we asked the model to predict the top three specialties that would handle the patient’s based on the symptoms and the patient info, for the general user and adding the initial vitals for the clinical user. More insights on the two evaluation frameworks and the two user scenarios can be read upon in the Methods: Specialty evaluation Framework.

In the first evaluation, which checked each of the top three predicted specialties individually if it matches the specialties in the ground truth, Claude 3.5 Sonnet slightly outperformed the other models. However, the performance differences among all models were minimal, with all models showing similar accuracy across both general and clinical user scenarios.

For the evaluation that focused on checking if at least one of the top three predicted specialties is predicted correctly, Claude 3.5 Sonnet had the highest performance, while overall performance differences remained small across all models. The results are illustrated in Figure 5 and Table 1.

**Figure 5:**
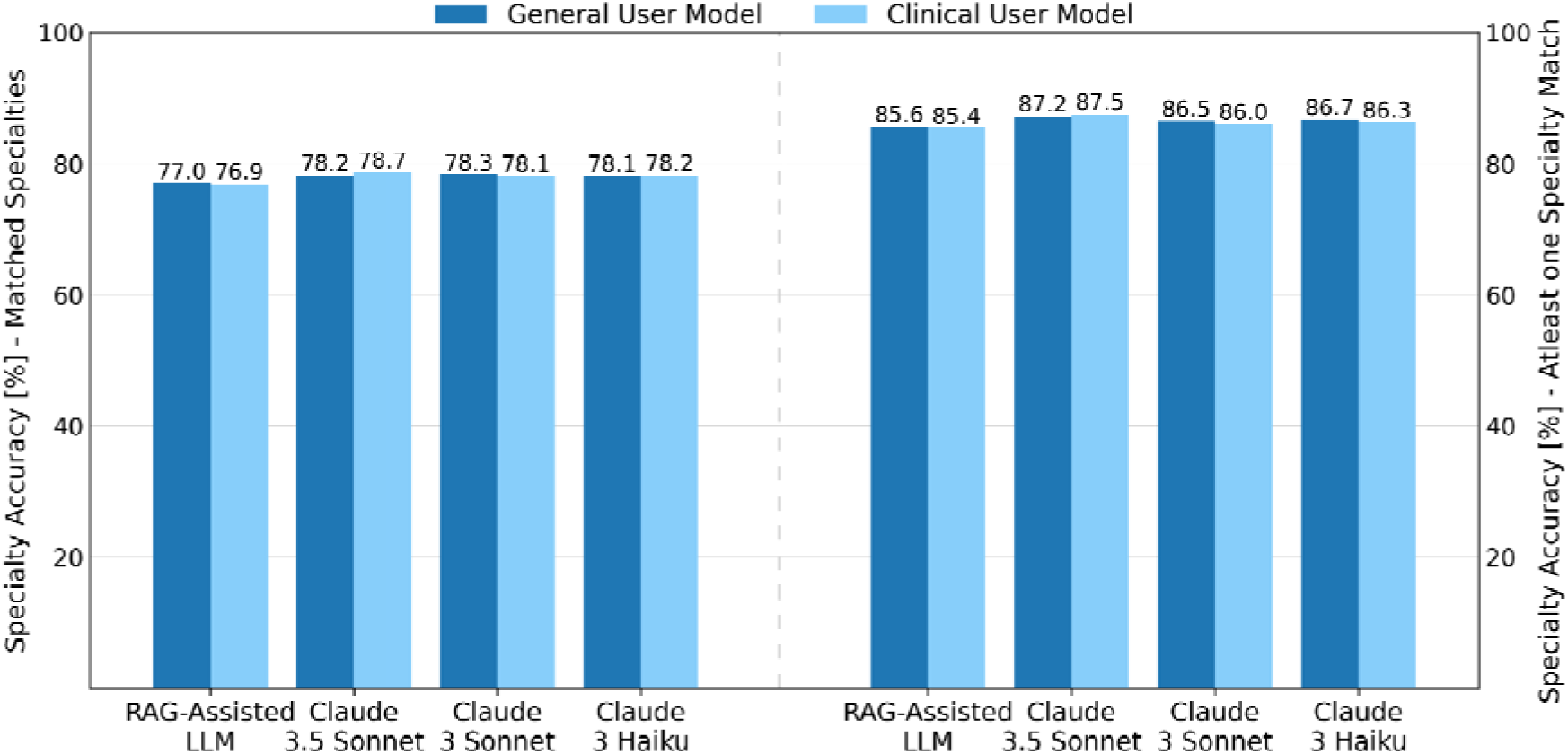
Performance as accuracy [%] on specialty with matched evaluation on the left and at least one diagnosis match evaluation on the right for both model types.

The improvement shown in Table 2 between the general user and clinician user models was most evident in the Claude 3.5 Sonnet model, with only minimal improvements seen in the RAG-assisted LLM and Claude 3 Haiku. In contrast, Claude 3 Sonnet experienced a negative impact when provided with additional information about the initial vitals. Predicting the appropriate specialty relies on several factors. Symptoms need to be clear and accurate, but it’s common for symptoms to fall under the expertise of multiple specialists, and often additional tests are required to narrow down the appropriate referral. In this study, the specialty was defined by the patient’s discharge primary diagnosis, meaning the diagnosis was made after several tests and possibly after days of observation. As a result, the addition of initial vitals may not significantly influence specialty prediction, as more detailed information becomes available only later in the patient’s care.

The evaluation of specialty frequencies, which can be found in the Supplementary Information in Figure S7 shows that in the best model, clinical user Claude 3.5 Sonnet, general surgery, emergency medicine, infectious diseases, and internal medicine are overrepresented, while the underrepresentation of orthopedics is nearly balanced by the higher occurrence of orthopedic surgery. The same tendencies can be seen in the other models.

The performance of LLMs in predicting the specialties shows that LLMs are generally well-suited to assist in medical referrals by offering a variety of relevant specialty options.

### Evaluating LLM workflows for diagnostic accuracy

In the process of clinical decision-making, we evaluated whether LLMs can assist in predicting the diagnosis or diagnoses a patient might have. We conducted this evaluation in our two settings like described before, the general user and clinical user setting. More on the evaluation framework can be found in the Methods: Diagnosis Evaluation Framework.

In our evaluation of LLMs’ ability to assist in predicting patient diagnoses, we found small differences in performance between models. In the first evaluation, in which each diagnosis was compared to the ground truth, Claude 3.5 Sonnet and Claude 3 Sonnet performed equally well for the general and clinical user setting. In the second evaluation, where the goal was to predict at least one correct diagnosis for each patient, all models demonstrated stronger performance. All results are presented in Figure 6 and in Table 1.

**Figure 6:**
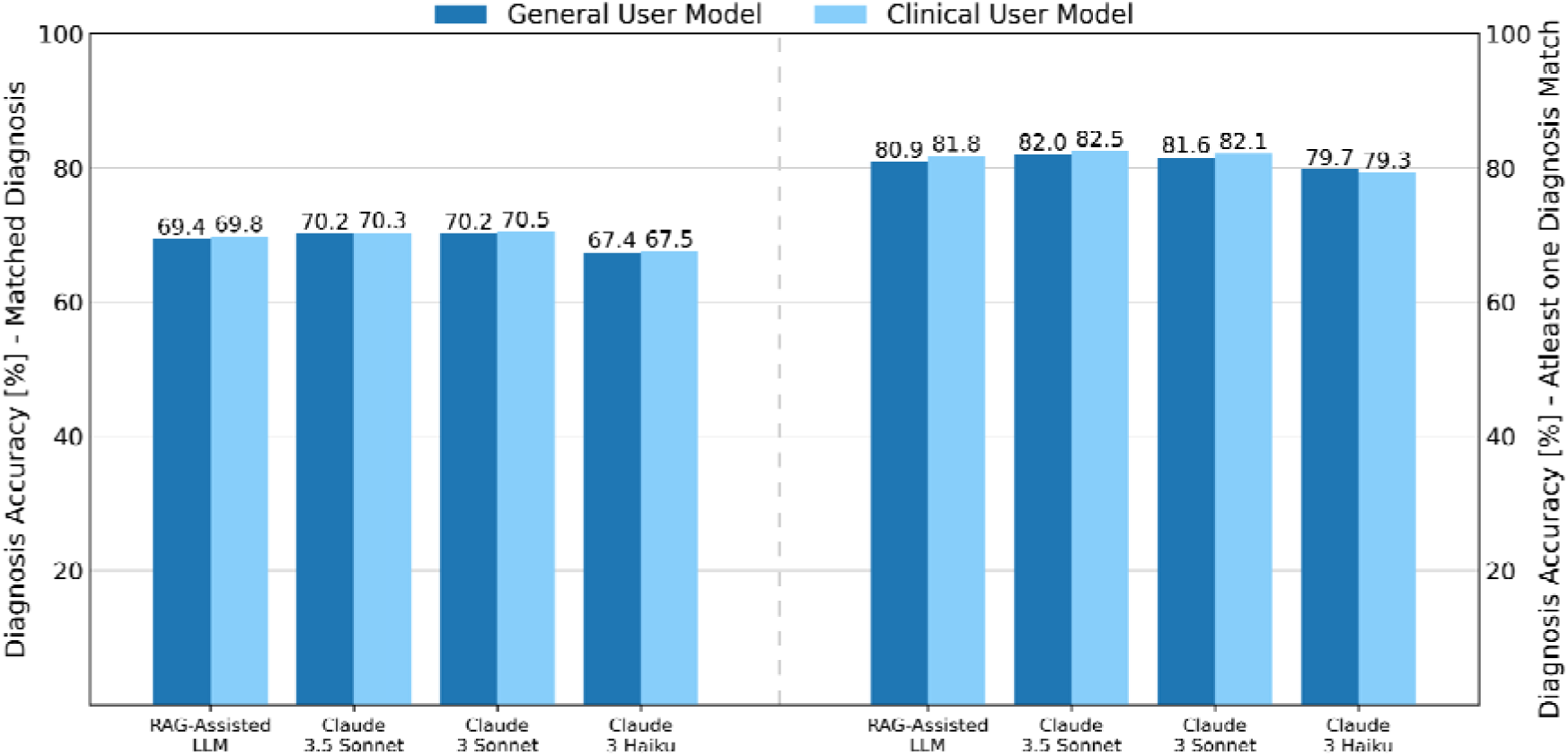
Performance as accuracy [%] on diagnosis with matched evaluation on the left and at least one diagnosis match evaluation on the right for both model types.

Improvements in the clinical user model over the general user model is particularly notable for the RAG-assisted LLM, as shown in Table 2. This suggests that the knowledge provided to the LLM during the RAG workflow has enhanced its diagnostic skills, particularly in interpreting and utilizing current initial vitals. Predicting or defining a diagnosis, like specialty referral, requires a significant amount of information, much of which is difficult to gather upon a patient’s arrival to the ED. This complexity underscores the challenges of early diagnosis in such fast-paced settings, where many crucial details are still emerging.

### Intra-Model Agreement

The agreement between the models can be seen as a measure of quality, as high agreement indicates that similar patterns and trends are captured in the responses, indicating robustness and reliability of the predictions. The analysis of inter-model agreement for the diagnosis data was omitted as this data has the highest variability and therefore requires assessment by an LLM judge.

Comparisons of inter-model agreement across the triage and specialty datasets are shown in Table 3, with full results available in Table 6. The intra-model agreement analysis showed the highest consistency between the general user model and the clinical model for all models. This suggests that the different inputs to the same model do not significantly alter or improve responses, but also that the models - particularly Claude 3.5 Sonnet and RAG-assisted LLM - show consistent performance across different user settings.

**Table 3:** Average inter-model agreement [%] for different categories over triage level and specialty. The “Between general user and clinical user” category shows the average agreement between the corresponding general user model and clinical user model, while the other categories show the average agreement to a certain model of the same type (general user to general user and clinical user to clinical user). Agreement to the same model is omitted to avoid distorting the average.

| Category | RAG-Assisted LLM | Claude 3.5 Sonnet | Claude 3 Sonnet | Claude 3 Haiku | Average |
| --- | --- | --- | --- | --- | --- |
| Between General and Clinical User | 85.94 | 89.74 | 84.95 | 83.54 | 86.04 |
| To RAG-Assisted LLM | - | 83.88 | 76.64 | 74.34 | 78.29 |
| To Claude 3.5 Sonnet | 83.88 | - | 77.19 | 73.35 | 78.14 |
| To Claude 3 Sonnet | 76.64 | 77.19 | - | 75.65 | 76.49 |
| To Claude 3 Haiku | 74.34 | 73.35 | 75.65 | - | 74.44 |

RAG-Assisted LLM demonstrated the highest average agreement across all models, closely followed by Claude 3.5 Sonnet, while the highest single inter-model agreement was between Claude 3.5 Sonnet and RAG-assisted LLM.

The high agreement between the models underlines their consistency in many cases, but the variation in agreement suggests that different models correctly classify different cases. This indicates that if we could determine which model excels at specific classifications, we could potentially reduce the overall error rate by a significant margin.

## Discussion

Advances in large language models (LLMs) are beginning to reshape how clinicians approach medical decision-making. These models have already proven useful in more structured tasks, like medical licensing exams, but how they can be used in real-world patient care is still being studied. We explored the potential of LLMs with and without RAG assistance, to support clinical decision-making by benchmarking their performance on 2000 real-world medical cases from the MIMIC-IV-ED dataset. We wanted to assess their ability to predict diagnoses, recommend specialists, and determine the urgency of care. Our results highlight both the promise and limitations of LLMs in the clinical decision process, offering insights into their potential role in healthcare.

Our results suggest that LLMs and the RAG-assisted LLM can support clinical decision-making, but their effectiveness varies depending on the task. Claude 3.5 Sonnet generally performed slightly better across most tasks, but the RAG-assisted LLM offered an important advantage: the ability to use external, trusted references. This feature helps reduce the risk of hallucinations and adds a layer of fact-checking, which is crucial in clinical settings where accuracy is crucial. The RAG-assisted LLM, compared to its base model Claude 3.5 Sonnet, showed a different pattern of improvement when using the clinical user setting (with additional patient vitals data), as demonstrated in Table 2. The RAG-assisted LLM benefited significantly from the extra vital information in the triage level and diagnosis tasks, though less so in the specialty task. In contrast, Claude 3.5 Sonnet showed improvements in the triage level and specialty tasks but gained less from the vital signs in the diagnosis task.

The RAG workflow allows the model to incorporate external sources from a research context, helping to provide a more informed perspective on the input. We hypothesize that the available external information likely emphasizes the relationship between vital signs and triage level or diagnosis, but not as much between vital signs and the corresponding referral specialty. Therefore, with the background knowledge provided by the RAG workflow, it makes sense that this model benefits more from additional vital signs in the domains of triage level and diagnosis. This also suggests that the RAG workflow improves the model’s performance in cases where current research findings are particularly relevant.

This is further highlighted by the RAG-assisted LLM’s strong performance in terms of exact accuracy on the triage level data with vital signs information, which is likely to be well-represented in available research resources. However, this does not necessarily help the model with range accuracy, as research sources are unlikely to guide the model in predicting more severe over less severe.

However, the benefit of incorporating more clinical information was not seen in simpler models like Claude 3 Haiku, and only minimal gains were observed for Claude 3 Sonnet when predicting specialties. This is in line with previous findings that LLMs struggle with nuanced clinical data, like interpreting abnormal lab results or subtle symptoms^8^. It also explains why none of the models achieved high accuracy in predicting the most severe triage patients, as these models are not equipped to follow numeric-based guidelines effectively^3^. More advanced models, like Claude 3.5, showed are better at handling these complexities.

A critical aspect of utilizing LLMs in clinical decision-making is the importance of prompt design. In our study, we experimented with various prompts to guide the models effectively, and it became evident that how a task is framed significantly impacts the quality of the results^27,28^. While we observed promising outcomes, it is clear that a more focused approach to prompt engineering would be highly beneficial, particularly when combined with the context of external sources provided by the RAG workflow. One interesting observation was the differences in performance between the LLM models. The models did not always agree on their predictions, which points to both a limitation and an opportunity. The results on intra-model agreement reveal that the models do not completely overlap in their predictions, suggesting that they might function as a “mixture of experts” when combined. Leveraging this diversity in predictions could lead to improved outcomes by utilizing the strengths of each model in different contexts. Additionally, higher agreement between models can be seen as a measure of quality, as it indicates that similar patterns and trends are being captured, contributing to the robustness and reliability of the predictions.

Finally, while our study establishes benchmark tasks and resources for clinical decision-making, the next step will involve refining the RAG-based model and similar approaches, and focusing on integrating them more effectively into clinician workflows. Beyond helping healthcare providers, these models can also benefit patients directly. For those experiencing symptoms at home, LLMs can provide an initial assessment, giving patients an indication for the severity of their condition and recommending which specialist to visit. This empowers patients to make more informed decisions about their care.

While there is still room for improvement, particularly in enhancing accuracy and reliability, our study suggests that LLMs already demonstrate relatively high response quality. As a result, they could play a significant role in streamlining clinical workflows and providing valuable health insights to patients. These advancements hold promise for faster, more accurate care and improved outcomes in a variety of healthcare settings.

## Methods

### Data Preprocessing

Our goal was to develop a model capable of predicting the specialty, triage level, and diagnosis for patients in an emergency department (ED) setting or those experiencing symptoms at home. Since we aimed to evaluate the difference in model performance based on whether the information was entered by the patient themselves or a clinician, we designed our dataset accordingly. For the general user, we required two main inputs: a description of the patient’s symptoms and some basic patient information. For the clinical user we added the initial vitals signs, such as temperature, heart rate, respiratory rate, oxygen saturation, and blood pressure, which can be measured upon arrival at the ED.

We processed and created our curated dataset using the MIMIC-IV ED dataset^22,25^ in conjunction with the MIMIC-IV Notes^22,26^ dataset, both modules from MIMIC-IV^20–22^, to support clinical decision-making in an emergency department setting. The MIMIC-IV ED dataset contains extensive information from patients admitted to the emergency department, while the Notes module provides valuable unstructured clinical notes of these patients. Specifically, we needed to extract symptoms, which were recorded in the form of “history of present illness” within the MIMIC-IV Notes “discharge” file as free text. Additionally, we extracted patient demographics such as gender, age, and race from the MIMIC-IV-ED “edstays” file, though age was extracted from the MIMIC-IV hospital data, where it is stored within the “patients” file. To complete the dataset, we required the patient’s initial vital signs, which were extracted from the MIMIC-IV-ED “vital signs” file. To compare our predictions, we also needed the triage level, which was retrieved from the MIMIC-IV-ED “triage” file. Furthermore, we needed the diagnoses, which were found in the form of “discharge primary diagnosis” in the MIMIC-IV Notes “Discharge” file as free text.

The preprocessing went as follows: Initially, we extracted relevant discharge notes from the IV Notes dataset and linked them with patient records using the *stay_id* from the MIMIC-IV ED 2.2 ED “stays” file. We filtered out duplicate *stay_id* entries to ensure the uniqueness of patient encounters. Triage information was then merged into the dataset, followed by the addition of patient demographic data, including gender and race, from the same “edstays” file.

Next, we incorporated diagnostic information and retained only records where the sequence number (*seq_num*) was equal to 1. Vital signs and other patient information were integrated, and we removed any records with missing triage levels.

For the clinical text data, we extracted only those entries that contained a history of present illness (HPI) within the raw discharge notes. The HPIs were filtered to ensure they were between 50 and 2000 characters in length, and further preprocessed to isolate symptom descriptions. We excluded any text containing the term “ED” (emergency department) and similar to maintain focus on the primary symptoms. Discharge diagnoses were also extracted from the raw discharge notes and preprocessed, separating primary from secondary diagnoses and discarding records that contained more than 15 primary diagnoses to maintain data consistency and relevance. This approach ensures that the dataset accurately reflects patient information and vital signs at the time of emergency department triage, offering a comprehensive view of early-stage clinical decision-making.

### Prompts

We created a series of prompts to guide the LLM in performing specific clinical tasks. These included predicting the triage level, predicting the specialty and diagnosis both together as they are both related and complement each other. Additionally, we used the prompt creating a ground truth referral specialist, and using the LLM as a judge to compare predicted diagnoses with the true diagnoses. Each prompt begins by setting the system’s role, such as, “You are an experienced healthcare professional with expertise in medical and clinical domains,” followed by clear task instructions. We also provided the data necessary for each task and specified how the LLM should format its responses, ensuring concise answers within predefined tags. The different prompts can be seen in the Supplementary Information in Figures S2 - S6.

### Model Selection

To comply with privacy regulations restricting the use of the MIMIC-IV dataset with external APIs like OpenAI’s GPT-4o and the Claude family models, we employed AWS Privatelink to securely connect to the Claude models hosted on AWS. This kind of evaluation reduces the likelihood that the data has been previously seen by the LLM models, which cannot be guaranteed when using publicly available datasets.

Claude 3.5 Sonnet, Claude 3 Sonnet, and Claude 3 Haiku are advanced LLMs developed to enhance natural language understanding, with improvements in performance and efficiency across multiple benchmarks over their predecessors, including GPT-4o, GPT-4T, Gemini 1.5 Pro and Llama 3 400B^23^. They excel in contextual understanding, efficiency, and their ability to handle specialized queries. This makes them well-suited for applications in clinical decision-making, where precision and adaptability are essential.

Claude 3 Haiku is the fastest and most compact model in Anthropic’s Claude 3 family. It excels in tasks where it requires quick analysis and response times^24^, making this feature suitable for the clinical-decision process.

Claude 3 Sonnet is a balanced combination of speed and intelligence, offering significant improvement in reasoning and accuracy. This model is versatile, handling complex text generation, analysis and reasoning^24^.

Claude 3.5 Sonnet is built on the foundations of Claude 3 Sonnet with further enhancement in speed and intelligence. It excels in different tasks like reasoning and question answering, while being faster and cost-efficient relative to the previous models. It has shown competitive or superior performance in a variety of language-based tasks^23^.

### Triage Level Evaluation Framework

The triage level is based on the Emergency Severity Index (ESI)^19^, which consists of five levels, as outlined in the Supplementary Information Table S1. We evaluate the model’s triage level predictions using two different assessment frameworks. The first is a straightforward comparison between the predicted triage level and the ground truth, with accuracy as the metric. The second evaluation framework uses a triage range approach, accounting for the variability in clinical judgment when assigning triage levels. The ESI is typically determined by a clinician assigning a score based on their assessment of a patient’s condition. Although there are defined levels within the ESI system, ranging from 1 to 5, the assignment of these levels can vary due to the clinician’s intuition and experience. In some cases, clinicians may lean on the side of caution, assigning a more severe level to avoid the risk of patient deterioration or the possibility of misclassifying a patient as less critical than they actually are. To account for this variability, our evaluation allows some flexibility in model predictions. If the real triage level value is 1, the model must predict 1, as immediate life-saving intervention is required. For a real value of 2, the model can predict either 1 or 2, ensuring patients needing urgent care aren’t harmed by overclassification. Similarly, if the real value is 3, the model can predict 2 or 3, and so on—up to a real value of 5, where the model can predict either 4 or 5.

### Specialty evaluation Framework

Since the MIMIC-IV-ED and MIMIC-IV-Notes datasets lack information on the medical specialist each patient visited, we used Claude 3.5 Sonnet to predict the most likely specialist for each diagnosis for each case, given that patients often present with multiple diagnoses rather than just one, thereby establishing the ground truth for this study.

Predicting a single specialist would be insufficient and unfair to the model when comparing its performance to the ground truth consisting of several specialties. In fact, it’s not uncommon for a patient to suffer from several medical conditions simultaneously, each requiring attention. To address this complexity, we chose to predict the top three specialists for each case. An Example is provided in Table 4.

**Table 4:** Example of a case with primary diagnoses, their corresponding created Ground Truth, and the predicted specialties for the case.

| Primary Diagnoses | Ground Truth Specialties<br>(Claude 3.5 Sonnet) | Predicted Specialties<br>(Claude 3.5 Sonnet) |
| --- | --- | --- |
| • Alcohol withdrawal<br>• Pancreatitis<br>• Thrombocytopenia<br>• Schizoaffective<br>• HIV | • Addiction Medicine<br>• Gastroenterology<br>• Hematology<br>• Psychiatry<br>• Infectious Disease | • Gastroenterology<br>• Hepatology<br>• Addiction Medicine |

This approach provides a more realistic comparison and offers clinicians and patients multiple possibilities to consider, reducing the risk of bias toward a single diagnosis. Ultimately, the LLM serves as a support tool, providing valuable insights, while the clinician makes the final, informed decision based on both the LLM’s recommendations and their own expertise.

### Diagnosis Evaluation Framework

As mentioned in the specialty evaluation previously, patients often come in with more than one diagnosis. To reflect this, we predicted a top three list of diagnoses for each case. We then compared each of these predictions to the actual diagnoses. To make the comparison more accurate, we used an LLM judge to decide if the predicted diagnosis either matched the ground truth or fit into a broader category of one of the actual diagnoses. This way, we accounted for differences in wording while still ensuring a fair evaluation.

We employed two evaluation methods for assessing the model’s performance in predicting the correct specialty. The first method evaluated whether each predicted specialty appeared in the ground truth list. For each patient, we counted how many specialties were correctly predicted and then divided that number by the length of the shorter list, either the ground truth or the prediction list.

For example, if the ground truth for a patient included only one entry, a cardiologist, and the model predicted three specialists—one cardiologist, one general medicine, and one electrophysiologist—only the cardiologist would be considered correct. Although general medicine and electrophysiology could also be relevant in some cases, our evaluation was specifically set to match the ground truth. This ties into a point discussed in the paper, where we explore how a single diagnosis might be managed by multiple specialists, a factor we plan to address in future work.

In this example, since only the cardiologist was correctly predicted, the patient would receive one point, which is then divided by the length of the shorter list (in this case, one, as the ground truth had only one entry). So, the score for this patient would be 1. If the ground truth had included two specialties, and the model only correctly predicted one out of three, the score would be 0.5. The total points across all patients were then summed and divided by the total number of patients to calculate the overall accuracy.

The second evaluation framework was simpler, focusing on whether at least one of the predicted specialties appeared in the ground truth list. If any one of the model’s predicted specialties matched one of the ground truth specialties, the prediction for that patient was considered successful.

### LLM Judge

For our study, we utilized LLMs to evaluate and compare the accuracy of predicted diagnoses for a given set of patient cases. This evaluation aimed to assess the model’s diagnostic capabilities by comparing the predicted diagnoses with those listed in the patient’s medical records. The prompt for the evaluation can be found in the Methods: Prompts.

The model was given the true list of diagnoses for each patient, along with three predicted diagnoses. It was then asked to determine if the predicted diagnosis matched the real one or if it fell under a broader category related to the real diagnosis. If it did, the model returned “True,” and if not, it returned “False”.

Similar methodologies have been explored successfully in recent research, showing that LLMs can effectively perform human-like evaluations in various tasks, including text summarization, quality assessments, and chat assistant evaluations, with results aligning closely to human judgments^29–31^. These findings support the use of LLMs as reliable tools for tasks like our diagnostic comparison evaluation.

While promising, the reliability and interpretability of LLMs as evaluation tools in real-world clinical environments still need further validation and refinement to ensure their safe and effective use.

### Intra-model agreement

We evaluated the agreement between models by comparing the predictions of different variants of the eight models, consisting of the RAG-assisted model and the three Claude language models with general user and clinical user settings each. Agreement was calculated separately for triage level predictions and specialty predictions and is symmetrical. Therefore, the results for both datasets are shown in in the Supplementary Information in Table S2, where the upper triangular matrix shows the intra-model agreement for triage and the lower triangular matrix for specialty, excluding self-comparisons (i.e., perfect agreement with the same model).

We evaluated and highlighted the two highest agreement values between model pairs for each dataset (specialty and triage) and for each of the three model user setting subgroups (general user to general user, general user to clinical user, clinical user to clinical user).

## Data Availability

The data will be available in pyhsioNet, we are in the process of submission. The core data set is available freely under https://physionet.org/content/mimic-iv-note/2.2/ and https://physionet.org/content/mimic-iv-ed/2.2/.

## Code Availability

The code to process data is available at https://github.com/BIMSBbioinfo/medLLMbenchmark

## Acknowledgments

We thank Akalin lab members for comments on the manuscript.

## Author contributions

AA conceptualized and planned the project. FG and MS jointly executed all of the computational analyses. FG made all the figures, completed benchmarks and wrote the initial draft of the manuscript. MS designed and implemented the RAG-based workflow. VF, MS, AA, and FG edited the manuscript. AA acquired funding and supervised the project.

